# Spread of the SARS-CoV-2 UK-variant in the South East of France: impact on COVID-19 patients’ age, comorbidity profiles and clinical presentation

**DOI:** 10.1101/2021.04.12.21253817

**Authors:** Johan Courjon, Julie Contenti, Elisa Demonchy, Jacques Levraut, Pascal Barbry, Géraldine Rios, Jean Dellamonica, David Chirio, Caroline Bonnefoy, Valérie Giordanengo, Michel Carles

## Abstract

**Objectives:** The variant 20I/501Y.V1, associated to a higher risk of transmissibility, emerged in Nice city (South East of France, French Riviera) during January 2021. The pandemic has resumed late December 2020 in this aera. A high incidence rate together with a fast turn-over of the main circulating variants, provided us the opportunity to analyze modifications in clinical profile and outcome traits.

**Methods:** Observational study in the University hospital of Nice from December 2020 to February 2021. We analyzed data of sequencing of SARS-CoV-2 from the sewage collector and PCR screening from all positive samples at the hospital. Then, we described the characteristics of all COVID-19 patients admitted in the emergency department (ED) (n=1247) and those hospitalized in the infectious diseases ward or ICU (n=232). Demographic data, clinical signs and severity were recorded by the NEWS-2, SAPS-2 and SOFA scores were recorded and analyzed.

**Results:** the UK-variant was absent in the area in December, then increasingly spread in January representing 59% of the PCR screening performed mid-February. The rate of patients over 65 years admitted to the ED decreased from 63% to 50% (p=0.001). The mean age of hospitalized patients in the infectious diseases ward decreased from 70.7 to 59.2 (p<0.001) while the proportion of patients without comorbidity increased from 16% to 42% (p=0.007). Neither the NEWS-2 score nor the main signs of clinical severity have changed over time.

**Conclusion:** Spread of the UK-variant in the South East of France affects younger and healthier patients.

## Introduction

Starting in September 2020, the new SARS-CoV-2 variants called VOC-202012/1 (lineage B.1.1.7, 20I/501Y.V1 or GR/501Y.V1) have rapidly spread in the UK, associated to a higher risk of transmissibility [1]. A matched cohort study including 109 812 patients in the UK showed an increase in deaths from 2.5 to 4.1 per 1 000 cases detected associated to the 20I/501Y.V1 variant [2].

The spread of this variant (20I/501Y.V1 *i*.*e*. “UK-variant”) in continental Europe was identified late December 2020, as reported by the European Center for Disease Prevention and Control (ECDC) with the first reports in Netherland, Belgium, Denmark [3]. It was reported in France late December 2020 [4,5].

After a second outbreak in November (with an incidence up to 501/100000 inhabitants), the pandemic declined in France in December 2020. At December 10^th^, the administrative area around Nice, French Riviera, in the South East of France, which totalizes more than 1 million inhabitants, exhibited a stable incidence of new positive cases at 169/100,000, i.e. a slightly higher value relative to the whole French territory. Contrary to the rest of France, the pandemic has resumed in this area since late December, reaching an incidence of 583/100,000 on February 13^th^ [5].

At the same time, the UK-variant, emerged in January in our area. The specific epidemiologic trends of COVID-19 epidemic in our area, i.e. a high incidence rate together with a fast turn-over of the main circulating variants, provided us the opportunity to analyze the UK-variant-related clinical profile and outcome traits.

## Method

We designed an observational study to assess the clinical profile and outcome of patients infected with the SARS-CoV-2, before and during the spread of the UK-variant. Patients admitted in the Emergency department (ED), in the Infectious Diseases department (IDD) and the ICU at Nice University Hospital (Cote d’Azur University UCA), France, were part of this study. Data were extracted from the hospital electronic database (n°410) of patient’s record, registered on clinicaltrial.gov NCT04779021, also from the hospital Virology Lab and the Institut de Pharmacologie Moléculaire et Cellulaire, UMR7275 CNRS/UNS (for sewage samples analysis).

In the way to quickly characterize the clinical consequences of the spread of these new variants, we designed a three-step approach.

### First step

*Spread of the UK-variant in our area and among hospitalized patients*. Data of the sequencing of SARS-CoV-2 identified from the sewage collector of the city of Nice (overall gathering) in December and January, and PCR screening from all positive samples analyzed at the hospital Virology Lab from newly admitted and diagnosed patients (starting late January 2021).

### Second step

*Concomitant epidemiology and clinical characteristics at hospital admission of COVID-19 overtime*, by an evaluation over a three-month period, from December 1^st^ 2020 to February 22^nd^ 2021, of all patients admitted in the ED for COVID-19. Demographic data, clinical severity as recorded by the NEWS-2 scoring system and the follow-up (ward admission *versus* ambulatory follow-up). Patients coming from Elderly Care facilities clusters or transferred from another hospital with a SARS-CoV-2 positive PCR were not included in the study.

### Third step

*Clinical severity and follow-up of hospitalized patients for a SARS-CoV-2 pneumonia*, in the IDD or ICU over three fortnights:

– before the spread of the UK-variant in our area,i.e. from December 7^th^ to 21^st^ 2020 (**Bef-F**),
– the two fortnights after starting a PCR screening from all positive samples analyzed at the hospital Virology Lab from newly admitted and diagnosed patients (late January 2021), i.e. from January 24^th^ to February 7^th^ and from February 8^th^ to 22^nd^, 2021 respectively **Aft1-F** and **Aft2-F**)

Demographic data, clinical signs and severity were recorded by the NEWS-2 (ED), SAPS-2 and SOFA scores (ICU), CT-Scan findings and the follow-up were recorded and analyzed.

We assessed the UK-variant spread in sewage through the Nice wastewater plant, which treats the sewage for a population of 390,000 inhabitants. The daily incoming volume varied from 97,000 m3 (in December 15^th^, 2020) to 128,000 (January 21^st^, 2021), due to rain at the end of January. Sequencing followed the protocol of the Artic consortium (https://www.protocols.io/view/ncov-2019-sequencing-protocol-v3-locost-bh42j8ye), data being analyzed with their pipeline (https://artic.network/ncov-2019/ncov2019-bioinformatics-sop.html). For each position associated with 20I/501Y.V1 lineage (https://cov-lineages.org/pangolin_tutorial.html) the fraction of 20I/501Y.V1 lineage reads relative to the total number of reads is assessed.

The virological assessment of positive PCR for SARS-CoV-2 variants at the Virology Lab was done as follow : SARS-CoV-2 positive samples were screened for the presence of the 20I/501Y.V1 lineage using either TaqPath COVID-19 RT-PCR (ThermoFisher, Illkirch-Graffenstaden, France) or ViroBOAR Spike 1.0 RT-PCR (Eurofins Biomnis, Lyon, France) kits, following manufacturer’s instructions. Identification of the 20I/501Y.V1 variant was further verified by sequencing by sequencing a random set of samples in which the reliability of the RT-PCR screening tests was checked.. Briefly, Spike protein regions covering Δ69/70 and Δ144 deletions, as well as the N501Y substitution, were amplified through RT-PCR, and amplicons were sequenced by the Sanger method.

Patients who were hospitalized with an identification of the UK-variant at the Virology Lab (see upper) were paired (1:2) with patients hospitalized during the three first weeks of December (variant-free period), on age and gender. News-2 score, time 1^st^ symptoms-ED admission and further hospital orientation (ICU versus medical ward) were the main evaluated criteria.

Data were expressed as mean ± standard deviation SD. Comparisons are done using Chi-2, Fisher exact-test for categorical variables and Student-t test, Kruskall Wallis test and Man Whitney U-test for continuous variables comparisons, if required. All data were analyzed using SPSS software^®^.

## Results

### 1. Timeline of UK-variant spreading

The survey in sewage of the city of Nice showed that the UK-variant represented 2,6%, 8,3% and 79,1% of all identified SARS-CoV2 strains in December 2020, January 2021 and February 2021 respectively. Consistent with these data, the random sequencing of positive PCR recorded from December 18^th^ to 26^th^ at Nice University hospital showed no identification of the UK-variant. This analysis was coordinated by the French National Center for Coronavirus (Institut des Agents Infectieux, CNR, HCL-Lyon) and was performed on 25% all the 73 positive patient’s samples isolated during this time period.

Finally, a systematic screening PCR to identified latent variants was scheduled starting late January, on governmental recommendation. At this time, all positive PCR done at the Nice Hospital were screened. The rate of UK-variant during Aft1-F was 20% of all positive PCR, then 59% during Aft2-F.

Altogether, these data suggested that the UK-variant was absent in our area in December 2020, then increasingly spread in January 2021.

### 2. Cohort profile of Covid-19 patients admitted in ED, overtime

From December 1^st^ to February 28^th^, 1247 patients were admitted for a suspected or diagnosed SARS-CoV-2 infection (Elderly Care Facilities people not included), aged 66.1 ± 17.9 [17-102] years, 54.0% male gender. Six patients (0.5%) died in the ED, while n=731 patients (58.6%) were hospitalized in a medical ward and n=109 (8.7%) in ICU; the remaining, n=401 (32.1%) were sent back home (ambulatory follow-up). Patients and clinical characteristics are reported (Table 1).

**Table 1.** Patients and clinical characteristics of <u>Covid-19 patients admitted in the emergency department</u> from December 1^st^ to February 28^th^

|  | December<br>n=211 | January<br>n=457 | February<br>n=579 | *p* |
| --- | --- | --- | --- | --- |
| Age years – mean (SD) | 68.0 (17) | 67.3 (17.7) | 64.4 (18.3)*,** | 0.006 |
| Patients over 65 years – n (%) | 133 (63) | 265 (58) | 290 (50)*** | 0.001 |
| Male gender – n (%) | 121 (57) | 244 (53) | 309 (53) | *ns* |
| Time 1<sup>st</sup> symptoms-ED– days (SD) | 6.4 (4.3) | 7.1 (4.9) | 7.5 (4.4)*** | <0.001 |
| ***Clinical signs*** |  |  |  |  |
| Respiratory rate/min. – mean (SD) | 22.0 (7.1) | 23.8 (9.3) | 24.4 (8.4)*** | 0.001 |
| Max Oxygen flow L/min – mean (SD) | 2.7 (3.6) | 3.2 (5.9) | 3.9 (6.9) | *ns* |
| Max Temperature °C – mean (SD) | 37.3 (0.9) | 37.3 (0.9) | 37.4 (1.2) | *ns* |
| Min SpO2 % – mean (SD) | 91.9 (6.1) | 91.9 (7.6) | 91.8 (7.2) | *ns* |
| NEWS-2 score – mean (SD) | 5.1 (3.6) | 5.1 (3.6) | 5.3 (3.5) | *ns* |

Post hoc analysis (Fischer exact-test or Mann Whitney U-test, if required): \* p<0.05 versus December \*\* p<0.05 versus January

Trends in patient’s age (overall) and hospital location after ED (following the initial medical assessment) over these 3-month period are reported (Figure 1).

**Figure 1 :**
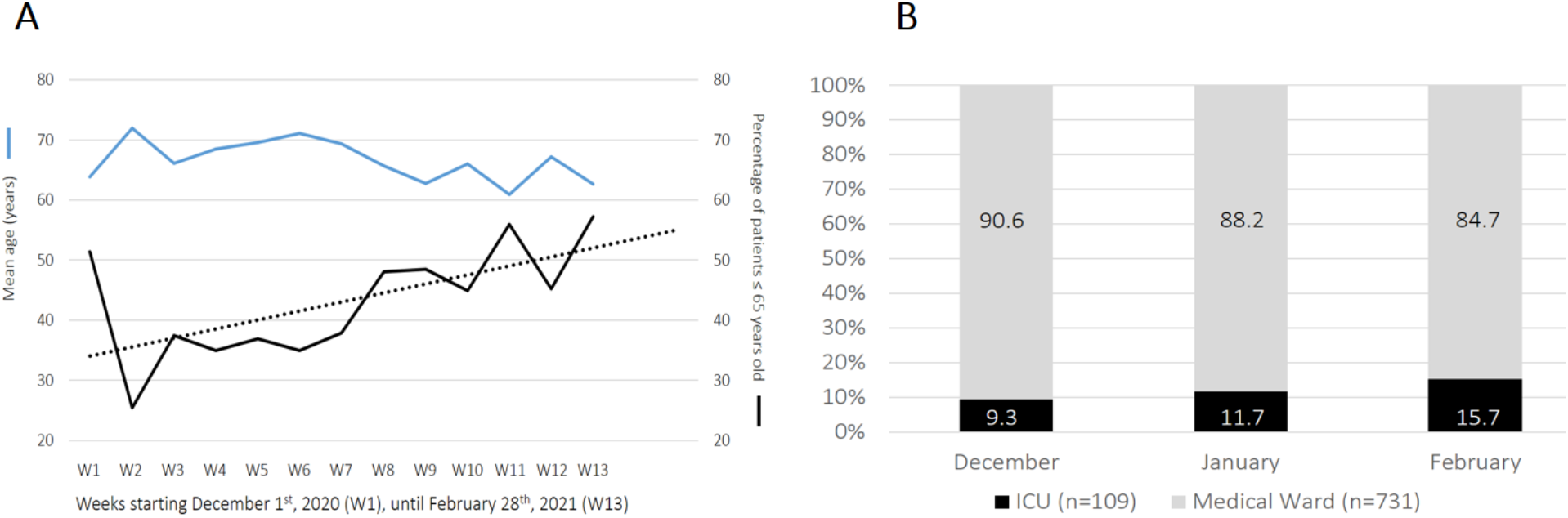
Age change and hospital admission after ED, over a 3 months-period. (**A**) mean age (blue line) and percentage of patients under 65 years old (black line, F-test 7.24, p=0.021) over weeks from December 1^st^, 2020, n=1247. (**B**) ICU *versus* medical ward admissions (100% normalized) for after ED hospitalization (n=840) over the three months period, February *versus* December, p=0.056.

Until February 22^nd^, we identified 29 hospitalized patients bearing the UK-variant. After pairing cases (n=29) *versus* controls (n=58) on age (61.0±10.7 vs 61.6±11.9 years) and gender (51.7% male in each group), we compared time 1^st^ symptoms-ED admission and the ED NEWS-2 score (cases vs controls) respectively: 7.3±3.5 vs 6.8±3.9, p=0.424 and 6.2±2.4 vs6.4±3.0, p=0.642. Patients admitted in the ED required immediate ICU management in respectively 10.3% (3/29) and 12.0% (7/58), p=0.810.

### Hospitalized patients in the Infectious Diseases and ICU departments : a three-periods assessment comparison

The three *a posteriori* defined groups, *i*.*e*. Bef-F, Aft1-F and Aft2-F respectively, included 51, 72 and 54 patients. Comparison of main data between groups are reported (Table 2).

**Table 2.** Hospitalized patients in the Infectious Diseases and ICU departments : a three-periods assessment comparison. Bef-F: December 7^th^ to 21^st^ 2020, Aft1-F: January 24^th^ to February 7^th^, Aft2-F: February 8^th^ to 22^nd^

| Infectious Diseases patients | Bef-F n=51 | Aft1-F n=72 | Aft2-F n=54 | p |
| --- | --- | --- | --- | --- |
| 1<sup>st</sup> symptoms - admission – days (SD) | 7.5 (4.5) | 6.9 (3.3) | 9.4 (5.6) ** | 0.01 |
| Age – mean (SD) | 70.7 (13.6) | 66.9 (15.8) | 59.2 (14.0) *,** | <0.001 |
| Male – n (%) | 33 (65%) | 44 (61%) | 31 (57%) | 0.745 |
| Diabetes – n (%) | 17 (33%) | 16 (22%) | 12 (22%) | 0.306 |
| Hypertension – n (%) | 20 (39%) | 30 (42%) | 15 (28%) | 0.252 |
| Obesity – n (%) | 10 (20%) | 18 (25%) | 9 (17%) | 0.503 |
| Immunosuppression – n (%) | 4 (8%) | 5 (7%) | 1 (2%) | 0.341 |
| No comorbidity – n (%) | 8 (16%) | 18 (25%) | 23 (42%) *,** | 0.007 |
| Charlson Index (CCI) – mean (SD) | 1.3 (1.6) | 0.9 (1.0) | 0.6 (0.8) *,** | 0.04 |
| **Clinical and radiological data** |  |  |  |  |
| Respiratory Rate/min – mean (SD) | 25.0 (6.1) | 26.9 (5.8) | 24.9 ± (5.9) | 0.051 |
| Room air SpO2% – mean (SD) | 90.5 (7.2) | 91.2 (5.6) | 92.1 (4.6) | 0.335 |
| CRP mg/L – mean (SD) | 94.3 (85.9) | 108.8 (77.7) | 94.4 (68.2) | 0.316 |
| CT-scan opacities<sup>#</sup> over 50% (n, %) | 8 (16%) | 10 (14 %) | 9 (17 %) | 0.951 |
| ICU admission | 12 (23%) | 19 (26%) | 19 (35%) | 0.374 |
| ICU patients | Bef-F n=13 | Aft1-F n=20 | Aft2-F n=22 | p |
| Age (years) – mean (SD) | 68.4 (10.8) | 64.5 (10.7) | 60.6 (12.5) | 0.154 |
| Male – n (%) | 9 (69%) | 13 (65%) | 16 (72%) | 0.863 |
| Mechanical ventilation – n (%) | 7 (54%) | 5 (25%) | 10 (45%) | 0.203 |
| HFNC – n (%) | 5 (38%) | 14 (70%) | 9 (45%) | 0.140 |
| SAPSII – mean (SD) | 40.6 (18.1) | 35.5 (15.3) | 40.0 (19.9) | 0.566 |
| Worst P/F – mean (SD) | 92 (33) | 112 (62) | 87 (24) | 0.244 |

<sup>#</sup> typical Covid-19 related imaging such as subpleural ground-glass opacities and subsequent consolidations, small vessel or intralobular septum thickening (33631941)
Note: Comparison between groups by a non parametric one way ANOVA on ranks (Kruskal Wallis test). Post hoc analysis (Fischer exact-test or Mann Whitney U-test, if required): \* p<0.05 versus December \*\* p<0.05 versus January

At the time of analysis, some data were not analyzed because of missing points due to shortness of follow-up with patients still hospitalized (length of stay, mortality).

In patients under 65 years old hospitalized in ID department (n=76), the rate of patients without any comorbidity rose from 21% (Bef-F) to 50% (Aft2-F), p=0.029.

## Discussion

The occurrence of UK-variant at the end of 2020 is of major concern, because of the contagiousness issue[1,4] and therefore the risk of an increased number of patients requiring an ICU bed. Beyond the good organization of the flow of an increased number of patients, the question arises as to whether this lineage is also associated with a more severe clinical presentation, which could involve younger patients. Our results show that patients admitted in January and February 2021 in ED for a COVID-19 and therefore hospitalized for a COVID-19-related pneumonia are significantly younger with around 50% of them aged less than 65 years old. Our data strongly support the increased risk of SARS-CoV-2-related severe pneumonia in younger patients contemporaneously to the spread of the UK-variant in our aera.

The severity issue is less conclusive. The rate of ED patients admitted in ICU increases, but an increased clinical severity is not obvious, as neither the NEWS-2 score nor the main signs of clinical severity (oxygen requirement, respiratory rate) have changed over time. Even when UK-variant patients were paired on age and gender, no difference in term of early stage severity was evidenced. In the same way, patients hospitalized in the ID department didn’t show a more severe inflammatory response or extended CT scan lung injuries at later times, i.e. after the spread of the UK-variant. Finally, for ICU patients, neither the severity score at admission (SAPSII) nor the depth of the respiratory distress seemed to increase by the variant. Altogether, there is no evidence to support a different and/or more severe clinical picture of the SARS-CoV-2-related pneumonia with the UK-variant.

The occurrence and severity of COVID-19 has been related to comorbidities like diabetes, hypertension or obesity [6,7]. Importantly, we provide herein data in favor of an increased ratio of healthy patients (i.e. without comorbidities) hospitalized for a SARS-CoV-2 related pneumonia associated to the spread of the UK-variant in the fifth-largest city of France. Combined to the patient’s younger age, this brings several concerns. The most immediate is about the risk of having a pandemic evolving toward young and healthy populations, with even greater social consequences. Another consequence, supported by our data is the increase of patients with younger age/comorbidity free-related ICU requirement. So far, older patients with comorbidities have a greater risk of ICU admission [8,9]. The increase of younger people admitted in ED for a COVID-19, in our cohort, is associated with a trend of increased ICU ratio requirement afterward. The same trend was noticed in the ID department. As the severity is not obviously increased, a likely explanation is that physicians, after months of fighting against the pandemic with critical patients mainly over 65 years old, have been surprised to receive younger patients with severe pneumonia, leading possibly to a higher ICU demand. As the pandemic rises concerns about ICU being overwhelmed, such change in patient’s profile brings new insights in the debate about ICU triage [10,11]. Furthermore, according to the country of interest, the vaccination strategy may have to be reconsidered in order to include a younger part of the population at risk of severe or critical COVID-19. Finally, the effect of the UK-variant on SARS-Cov-2-related mortality cannot be appropriately assessed here because, giving a mortality rate under 0.5%, the required sample size needs to be thousands of patients [2]. The number of patients managed in our hospital and the duration of follow-up available do not allowed us to consider the final outcome for comparison between study periods.

Until today, reports of a relationship between SARS-CoV-2 genetic specificities among variants and clinical presentation are scarce. One variant with a deletion (Δ382) in the open reading frame 8 have been associated to milder infections in Singapore[12] and may have play a role in the very low case fatality rate in this country [13]. Spike mutation D614G that probably occurs in China before its diffusion in Europe is associated with a decreased age of COVID-19 patients possibly due to an increased viral load in younger patients [14]. The N501Y mutation is associated with adaptation to rodent, for instance mice [15] and may increase SARS-CoV-2 Spike protein binding to ACE2 because of conformational changes, thus increasing its transmissibility [1]. The role of Δ69/Δ70 on the spike protein is also potentially involved in the increased transmissibility [16].

The spread of the UK-variant was found associated with a higher transmissibility as reported by others[4]. Since our data are mainly on hospitalized patients, the carriage of the UK-variant in a younger population bears further studies about the clinical fraction associated to the variant in the population [17] outside the hospital. A puzzling question is why younger patients need now to be hospitalized, which was not the case before? The higher transmissibility of UK-variant is supposed to be similar among all age groups [18]. As of 26^th^ of February, 60 000 inhabitants of Alpes Maritimes had received 2 doses of vaccine. Increased herd immunity after vaccination of the old community and mortality during the two previous epidemic peaks in France that has affected the most fragile elderly may have independently contributed to the observation of a decreased number of patients over 75 year-old requiring hospitalization.

Our study has some limitations. First, data are retrospective. Then, data on the UK-variant spread results from analysis of sewage or selected hospital samples and not from a systematic survey. Data on hospitalized patients (variant *versus* “common” strain) are incomplete. Finally, patient’s data were extracted from ED, ID and a medical ICU of the University hospital of Nice, but patients hospitalized in other hospitals from the ED were not included in the study, leading to incomplete analysis of involved patients. Our study do not allow to distinguish an effect of the UK-variant at the individual level, that is a higher probability to develop pneumonia after a contact with the virus due to the Spike protein RBD mutations, from a modification of virus diffusion in the South East of France population leading to a higher exposure of younger inhabitants which could be related to the virus itself or to a modification in non-pharmaceutical interventions adherence.

In summary, our data alerts on the consequences of the UK-variant spreading with younger and healthier patients requiring hospitalization for a SARS-CoV-2 pneumonia. Health system impact (with the risk of overwhelming ICU and medical units) as well as social consequences should be assessed in larger prospective studies.

## Data Availability

Data are available on request

## Acknowledgments

We thank Rainer Waldmann for fruitful discussions

